# Disparities in Access to Vascular Stroke Imaging and Carotid Revascularization: A Population Study

**DOI:** 10.1101/2025.03.26.25324731

**Authors:** Harshil Shah, Tina He, Naomi Dyck, Jillian Stang, Dana Nicol, Christiane Mcintosh, Stephen Wilton, Shelagh B. Coutts, Nishita Singh, Michael D. Hill, Aravind Ganesh

## Abstract

**Background:** CT angiography (CTA), MR angiography (MRA), and ultrasound are noninvasive vascular imaging modalities used in the investigation of stroke or transient ischemic attack (TIA). Imaging decisions may be influenced by factors ranging from location-based resource considerations to patient characteristics. The aim of this study was to investigate disparities in vascular imaging utilization and subsequent carotid revascularization over 7 years in a Canadian province (Alberta, population:4.4 million).

**Methods:** We used provincial administrative data encompassing patients presenting to hospital or emergency/urgent-care facilities with a diagnosis of TIA or ischemic stroke from 1-April-2016 to 31-Mar-2023 and related the vascular imaging received (CTA/MRA/ultrasound/none) to age, sex, region (rural vs urban), diagnosis (ischemic stroke vs minor stroke/TIA), comorbidities, center type, and year using multivariable logistic regressions. We explored whether these variations persisted in recurrent events and investigated the odds of carotid endarterectomy/stenting using similar regression models.

**Results:** Among 47,963 patients (median age: 72, interquartile range: 21, 47.6% female) with TIA/stroke, those older than seventy-one, with minor stroke/TIA, and with specific comorbidities had significantly lower odds of receiving CTA or any neurovascular imaging, as were those in rural sites or hospitals not designated as Comprehensive Stroke Centers (CSCs, e.g. aOR-CTA [stroke unit-equivalent care vs CSC]: 0.20, 95%CI:0.13-0.30). Female patients were less likely to undergo CTA or any vascular imaging (66.4% female vs 71.1% male, aOR:0.84, 95%CI:0.81-0.88). Those presenting in more recent years had higher odds of receiving CTA (aOR-per-additional-year:1.15, 95%CI:1.14-1.17) or any neurovascular imaging (aOR:1.13, 95%CI:1.11-1.14). Female sex was associated with lower odds of carotid revascularization, as were patients with minor stroke/TIA, atrial fibrillation, care at non-CSC centers, and absence of vascular imaging (e.g. aOR[female vs male]:0.57, 95%CI:0.51-0.64).

**Conclusions:** We found important demographic and geographic disparities in vascular imaging utilization despite increasing utilization over time; similar disparities were also seen in carotid revascularization.

## Introduction

Up to 20% of stroke and TIA events are attributed to large artery atherosclerotic stenosis of the common or internal carotid arteries, after which there is a high risk of recurrent stroke due to the unstable atherosclerotic plaque^1^. Carotid revascularization procedures, either stenting or endarterectomy, are efficacious in secondary prevention, especially if done urgently^2^. Vascular imaging of the head and neck plays a crucial role in establishing the presence and extent of contributory extracranial or intracranial vessel disease (most commonly atherosclerosis), thereby informing decisions on carotid procedures and medical therapies like antithrombotic regimens and statins^3^.

There are three noninvasive vascular imaging modalities that are used in stroke imaging: computed tomography angiography (CTA), ultrasound, and magnetic resonance angiography (MRA). However, across the world, there has been a general preference for CT angiography^4^, perhaps because the lumen diameter is well shown and best approximates digital subtraction angiographic images. Other advantages include relative accessibility and speed of completion in the acute setting, as well as acceptable quality of imaging for key parenchymal and extra/intracranial vascular structures^5^. Although CTAs are often the preferred vascular imaging in stroke, there is variation in use based on the center type and geography (urban or rural)^6^.

Disparities among regions may also extend to access subsequent carotid procedures. A Canadian study using discharge data from 1994-1997 revealed marked regional variation in carotid endarterectomy rates, attributed at the time to the differing views of indications and inconsistency of published guidelines^7^. As indications and guidelines become consistent and well-known, regional variations may persist due to resource limitations^8^.

Patient characteristics, such as race and sex, are associated with disparities in imaging following stroke/TIA and carotid revascularization. In the United States of America, Black patients receive diagnostic imaging less often than White patients after ischemic stroke^9^, and female patients are less likely to receive carotid revascularization procedures^10^. However, these prior studies were not population-based and may suffer from selection bias and other confounding factors.

We examined whole population-level disparities in receipt of vascular stroke imaging and subsequent carotid revascularization based on demographic and geographic factors among all patients presenting with stroke/TIA in the province of Alberta, Canada.

## Methods

### Data Source

This study used population-level administrative data for Alberta, a Western Canadian province with a population of 4.4 million^11^. We obtained data for all patients with ischemic strokes or TIA from April 1, 2016, to March 31, 2023 (N=47,963). This was derived from the Discharge Abstract Database, maintained by Alberta Health, for patients admitted to hospital, and the National Ambulatory Care Reporting System, for patients seen at emergency departments or other care centers who were not hospitalized. These databases use standardized definitions of ischemic stroke and TIA or minor stroke (i.e., patients presenting with ischemic stroke who were discharged home directly from the emergency department) based on the International Classification of Disease 10, which are 92-97% accurate^12,13^. The use of imaging and imaging type was recorded for each episode of care and enumerated. We used the Canadian Classification of Health Interventions codes to identify all carotid revascularization procedures or recurrent episodes which occurred until March 31, 2024.

### Analysis

The vascular imaging modality during the index event for each patient received was determined (MRA vs. CTA vs. ultrasound). CTA/MRA was enumerated if patients received this imaging modality in addition to ultrasound. Next, the type of center (comprehensive, primary, stroke equivalent, other) and area type (urban or rural) was classified for the 107 hospitals in the province using the 2016 census information which provides a list of urban and rural communities.

Univariable analyses were done for categorical variables using a chi-squared test, with age was dichotomized at the median. We also examined the association of the year of stroke with CTA, vascular imaging, and carotid procedure use using logistic regression. Multivariable logistic regression models were used to estimate the effect size of the association between use of CTA or any vascular imaging for both the index and subsequent events and key demographic and health service factors. All effect size estimates were adjusted for year, age, sex, region type, index event diagnosis, comorbidities, and center type. A similar regression model was conducted to investigate the effect size of the association between receiving carotid procedures and the abovementioned variables. Finally, this multivariable logistic regression was repeated to also adjust for imaging modality received. For this study, the STROBE guidelines for reporting observational studies were used^14^.

### Ethics Approval

The University of Calgary Conjoint Health Research Ethics Board (REB22-0233) approved this study and waived the need for informed consent for this population analysis.

## Results

This study consisted of 47,963 patients (median age: seventy-two, interquartile range: 61-82, 22,809 [47.6%] female) with 22,853 (47.6%) presenting with TIA/minor stroke (47.6%) and 25,110 (52.4%) presenting with ischemic strokes (Table 1). 40,123 (83.7%) of them presented to urban hospitals, with 17,891 (37.3%) of patients presenting to Comprehensive Stroke Centers. 29,563 (61.7%) of the patients had CTA as their most sophisticated vascular imaging, with 14,922 (31.1%) receiving no vascular imaging. 494 (1.0%) of patients received MRA, of which 262 (53.0%), also had CTA imaging. A total of 1638 (3.4%) patients received carotid revascularization. The median follow-up time was 57 months.

**Table 1:** Characteristics of patients with stroke or TIA between 2016 and 2023 in Alberta, Canada. (n=47963)

|  |  |  |  |  |  |  |  |  |  |
| --- | --- | --- | --- | --- | --- | --- | --- | --- | --- |
|  | 2016<br>(N = 6026) | 2017<br>(N= 7616) | 2018<br>(N=7299) | 2019<br>(N=7067) | 2020<br>(N=6874) | 2021<br>(N=6901) | 2022<br>(N=5325) | 2023<br>(N=807)* | Total<br>(N=47963) |
| Demographic |  |  |  |  |  |  |  |  |  |
| Median Age, yr, (IQR) | 71 (60-82) | 72 (60-82) | 71 (60-82) | 72 (61-81) | 72 (61-82) | 71 (61-81) | 72 (62-82) | 73 (63-82) | 72 (61-82) |
| Older Patients, >=72 years, N (%) | 2998 (49.8) | 3853 (50.6) | 3627 (49.7) | 3559 (50.4) | 3437 (50.0) | 3372 (48.9) | 2774 (52.1) | 447 (55.4) | 24088 (50.2) |
| Female Sex, N (%) | 2928 (48.6) | 3695 (48.5) | 3513 (48.1) | 3335 (47.2) | 3242 (47.2) | 3237 (46.9) | 2457 (46.1) | 376 (46.6) | 22809 (47.6) |
| Urban Region, N (%) | 4884 (81.0) | 6213 (81.6) | 6048 (82.9) | 5923 (83.8) | 5766 (83.9) | 5827 (84.4) | 4674 (87.8) | 745 (92.3) | 40123 (83.7) |
| Index Event Diagnosis |  |  |  |  |  |  |  |  |  |
| Ischemic Stroke, N (%) | 2890 (48.0) | 3550 (46.6) | 3437 (47.1) | 3521 (49.8) | 3548 (51.6) | 3674 (53.2) | 3746 (70.3) | 699 (86.6) | 25110 (52.4) |
| Minor Stroke/TIA, N (%) | 3136 (52.0) | 4066 (53.4) | 3862 (52.9) | 3546 (50.2) | 3326 (48.4) | 3227 (46.8) | 1579 (29.7) | 108 (13.4) | 22853 (47.6) |
| Comorbidities |  |  |  |  |  |  |  |  |  |
| Atrial Fibrillation, N (%) | 1536 (25.5) | 1803 (23.7) | 1575 (21.6) | 1341 (19.0) | 1221 (17.8) | 1075 (15.6) | 902 (16.9) | 122 (15.1) | 9594 (20.0) |
| Coronary Artery Disease, N (%) | 811 (13.5) | 944 (12.4) | 845 (11.6) | 708 (10.0) | 579 (8.4) | 560 (8.1) | 326 (6.1) | 49 (6.1) | 4835 (10.1) |
| Chronic Renal Failure, N (%) | 563 (9.3) | 691 (9.1) | 543 (7.4) | 446 (6.3) | 411 (6.0) | 410 (5.9) | 252 (4.7) | 32 (4.0) | 3354 (7.0) |
| Diabetes Mellitus, N (%) | 2025 (33.6) | 2381 (31.3) | 2210 (30.3) | 2281 (32.3) | 2119 (30.8) | 2056 (29.8) | 1705 (32.0) | 281 (34.8) | 15081 (31.4) |
| Hypertension, N (%) | 4133 (68.6) | 5180 (68.0) | 4736 (64.9) | 4383 (62.0) | 4348 (63.3) | 4235 (61.4) | 3805 (71.5) | 619 (76.7) | 31477 (65.6) |
| Heart Failure, N (%) | 1183 (19.6) | 1390 (18.3) | 1215 (16.6) | 1031 (14.6) | 913 (13.3) | 844 (12.2) | 675 (12.7) | 97 (12.0) | 7361 (15.3) |
| Dialysis, N (%) | 123 (2.0) | 136 (1.8) | 122 (1.7) | 112 (1.6) | 96 (1.4) | 113 (1.6) | 73 (1.4) | 10 (1.2) | 787 (1.6) |
| **Center Type** |  |  |  |  |  |  |  |  |  |
| Comprehensive Stroke Center, N (%) | 2130 (35.3) | 2636 (34.6) | 2648 (36.3) | 2541 (36.0) | 2596 (37.8) | 2667 (38.6) | 2282 (42.9) | 373 (46.2) | 17891 (37.3) |
| Primary Stroke Center, N (%) | 1666 (27.6) | 2005 (26.3) | 1917 (26.3) | 1927 (27.3) | 1901 (27.7) | 1849 (26.8) | 1475 (27.7) | 235 (29.1) | 12995 (27.1) |
| Stroke Unit Equivalent Care, N (%) | 16 (0.3) | 26 (0.3) | 34 (0.5) | 23 (0.3) | 17 (0.2) | 19 (0.3) | 16 (0.3) | 2 (0.2) | 153 (0.3) |
| None of the above, N (%) | 2214 (36.7) | 2949 (38.7) | 2700 (37.0) | 2576 (36.5) | 2360 (34.3) | 2366 (34.3) | 1552 (29.1) | 197 (24.4) | 16924 (35.3) |
| **Most Sophisticated Imaging, N (%)** |  |  |  |  |  |  |  |  |  |
| MRA/CTA | 2956 (49.1) | 4017 (52.7) | 4304 (59.0) | 4416 (62.5) | 4611 (67.1) | 4779 (69.3) | 3826 (71.9) | 654 (81.04) | 29563 (61.7) |
| Ultrasound | 628 (10.4) | 695 (9.1) | 583 (8.0) | 468 (6.6) | 369 (5.4) | 333 (4.8) | 329 (6.2) | 40 (5.0) | 3457 (7.2) |
| None of the above | 2442 (40.5) | 2904 (38.1) | 2412 (33.0) | 2183 (30.9) | 1894 (27.6) | 1789 (25.9) | 1170 (22.0) | 113 (14.0) | 14922 (31.1) |
| **Carotid Procedure, N (%)** |  |  |  |  |  |  |  |  |  |
| Any Carotid | 181(3.0) | 232(3.0) | 244(3.3) | 235(3.3) | 261(3.8) | 228(3.3) | 223(4.2) | 33(4.1) | 1638(3.4) |
| Procedure |  |  |  |  |  |  |  |  |  |

Note: IQR=interquartile range, TIA=transient ischemic attack, MRA=Magnetic Resonance Angiography, CTA=Computed
Tomography Angiography
\* There are fewer cases in 2023 as the cohort for index events concluded 31-March-2023

Over the years, there was a general trend of increased utilization of vascular imaging for patients with stroke/TIA (p_het_<0.001), with the most sophisticated vascular imaging being CTA more commonly and ultrasound less commonly (Figure 1).

**Figure 1.**
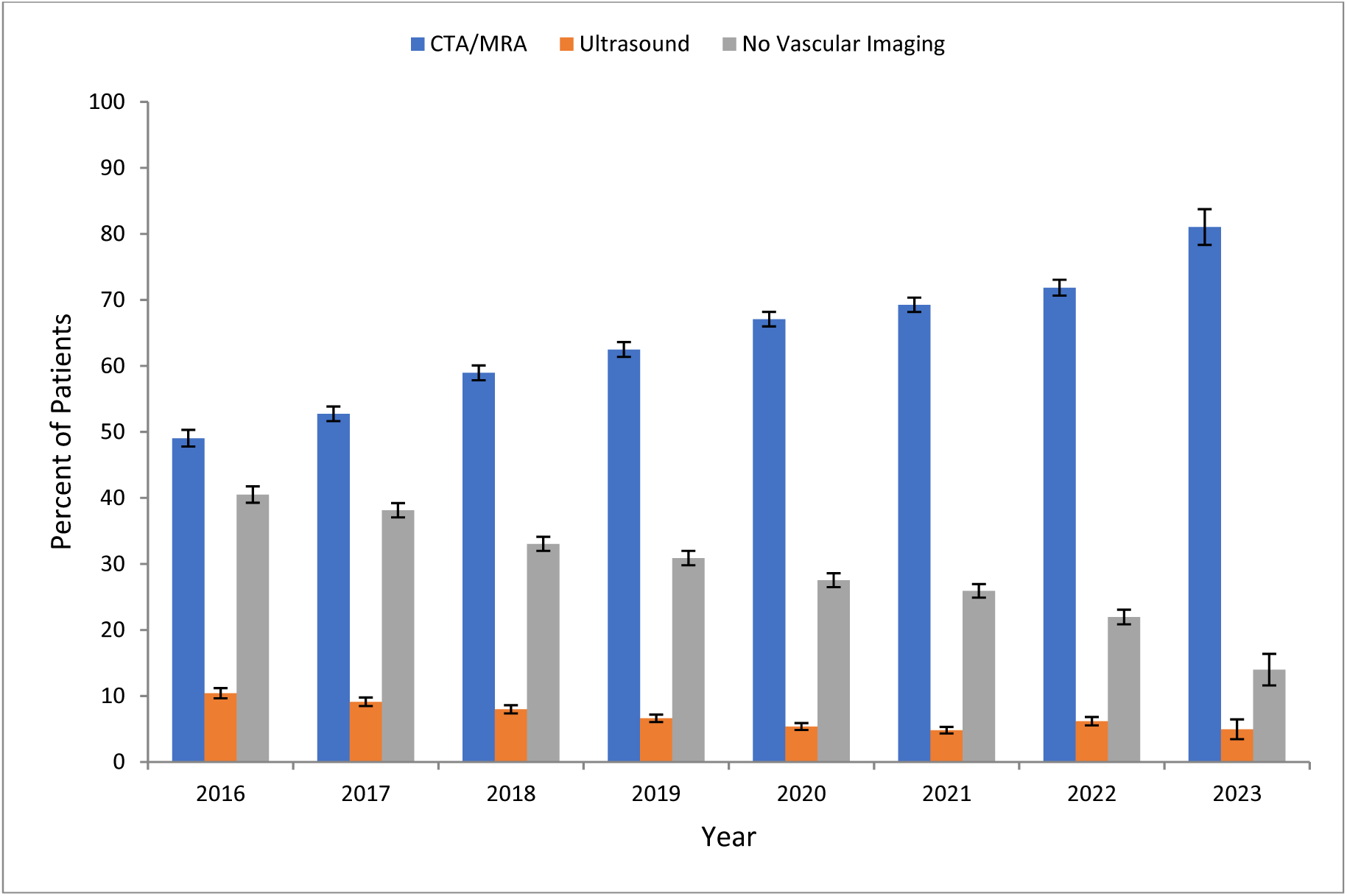
Bar chart depicting the most sophisticated imaging modality used as a percentage of cases each year from 2016 to 2023

Univariable analyses revealed female patients were less likely to receive CTA (58.2% of females vs 63.9% male, p_het_<0.001) or any vascular imaging (66.4% female vs 71.1% male, p_het_<0.001). Further univariable analyses also showed significant heterogeneity in receiving CTA and vascular imaging across age groups, sex, urban vs rural regions, index event diagnosis, and center type (p_het_<0.001). These differences generally persisted across all seven years, particularly for rural vs urban regions (Figure 2). Aside from age, these variables, in addition to primary imaging modality, were also significantly associated with receiving carotid revascularization procedures (p_het_<0.001). Patients with atrial fibrillation, coronary artery disease, chronic renal failure, diabetes, and hypertension, also showed significant heterogeneity in receiving carotid revascularization procedures.

**Figure 2.**
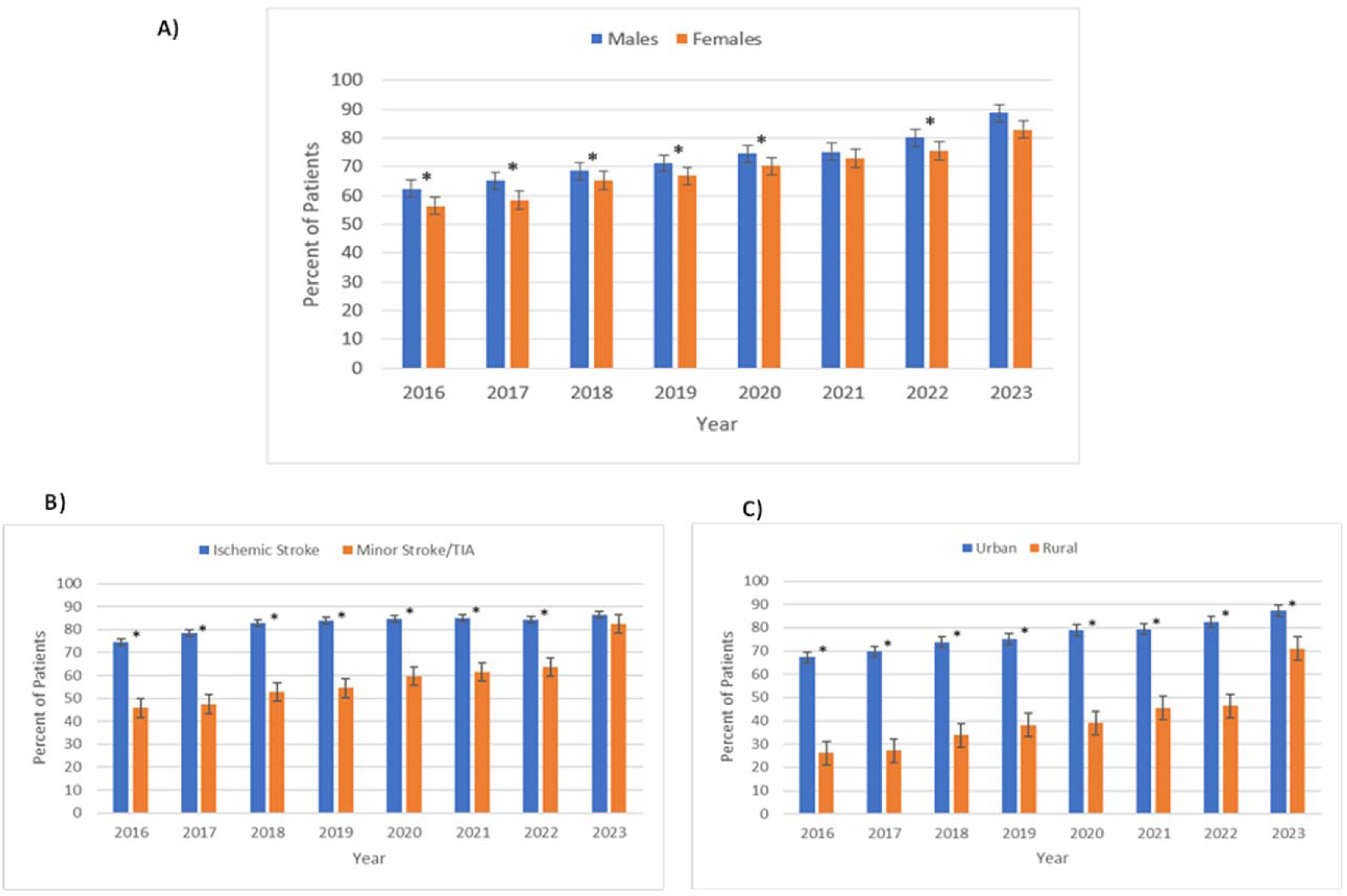
Bar graph depicting the percent of patients receiving any vascular imaging over the years separated by (A) sex, (B) ischemic stroke vs TIA/minor stroke indication, and (Cf) rural vs urban location. Asterisks represent 95% CI not overlapping, demonstrating a significant difference.

Upon examining the utilization of CTAs using multivariable regressions, we confirmed that odds of receiving CTA imaging increased over time (aOR per year since 2015:1.15, 95%CI:1.14-1.17). Further, patients who were female (aOR:0.83, 95%CI:0.79-0.86) or older than 71 (aOR:0.66, 95%CI:0.63-0.69) were less likely to receive CTAs, as were patients who presented to rural sites (aOR:0.48, 95%CI:0.46-0.51) versus urban sites. Those with minor stroke/TIA as the diagnosis also had lower odds of receiving CTA (aOR:0.51, 95%CI:0.49-0.53). Patients with chronic renal failure (aOR:0.59, 95%CI:0.55-0.65), dialysis (aOR:0.40, 95%CI:0.34-0.47), heart failure (aOR:0.80, 95%CI:0.75-0.85), diabetes (aOR:0.92, 95%CI:0.88-0.96), and coronary artery disease (aOR:0.89, 95%CI:0.83-0.95) were less likely to receive CTAs. Conversely, patients with hypertension (aOR:1.22, 95%CI:1.17-1.28) were more likely to receive CTAs. Finally, patients seen in Primary Stroke Centers (aOR:0.34, 95%CI:0.32-0.36), Stroke Unit Equivalent Centers (aOR:0.13, 95%CI:0.09-0.20), or other centers (aOR:0.21, 95%CI:0.20-0.23) were less likely to receive CTA than those seen at Comprehensive Stroke Centers (Table 2). When the receipt of any vascular imaging (regardless of modality) was examined, multivariable regression models revealed patients with diabetes and coronary artery disease did not have any significant differences in odds of receiving imaging. Otherwise, all other results were similar to those of receiving CTA (Table 2).

**Table 2:** Odds of vascular imaging utilization for index stroke/TIA from multivariable logistic regression.

| Variable | CTA Imaging | Any Vascular Imaging |
| --- | --- | --- |
| Years Since 2015 (per year) | 1.15 (1.14-1.17)* | 1.13 (1.11-1.14)* |
| Older Patients ( ≥72 years) | 0.66 (0.63-0.69)* | 0.73 (0.70-0.77)* |
| Female Sex | 0.83 (0.79-0.86)* | 0.84 (0.81-0.88)* |
| **Region** |  |  |
| Urban | Reference | Reference |
| Rural | 0.48 (0.46-0.51)* | 0.33 (0.31-0.35)* |
| **Indications** |  |  |
| Ischemic Stroke | Reference | Reference |
| Minor Stroke/TIA | 0.51 (0.49-0.53)* | 0.37 (0.35-0.38)* |
| **Comorbidities** |  |  |
| Atrial Fibrillation | 1.00 (0.94-1.06) | 1.03 (0.97-1.09) |
| Chronic Renal Failure | 0.59 (0.55-0.65)* | 0.75 (0.69-0.82)* |
| Dialysis | 0.40 (0.34-0.47)* | 0.51 (0.43-0.60)* |
| Diabetes Mellitus | 0.92 (0.88-0.96)* | 0.99 (0.94-1.04) |
| Heart Failure | 0.80 (0.75-0.85)* | 0.81 (0.76-0.87)* |
| Hypertension | 1.22 (1.17-1.28)* | 1.33 (1.27-1.40)* |
| Coronary Artery Disease | 0.89 (0.83-0.95)* | 0.95 (0.88-1.02) |
| **Center Type** |  |  |
| Comprehensive Stroke Center | Reference | Reference |
| Primary Stroke Centre | 0.34(0.32-0.36)* | 0.54 (0.51-0.58)* |
| Stroke Unit Equivalent Care | 0.13(0.09-0.20)* | 0.20 (0.13-0.30)* |
| None of the above | 0.21(0.20-0.23)* | 0.35 (0.33-0.38)* |

During follow-up period (up to March 31, 2024), 6208 patients in our study’s cohort had recurrent events. Similar logistic regression models were created for this population (Table 3). The odds of receiving CTA for recurrent events increased over time (aOR per year since 2015: 1.12, 95%CI:1.09-1.16) and any vascular imaging (aOR: 1.08, 95%CI:1.05-1.12). Older age (aOR: 0.81, 95%CI:0.72-0.92) decreased the odds of CTA but not vascular imaging. Female patients continued to have reduced odds of both CTA (aOR: 0.73, 95%CI:0.65-0.82) and any vascular imaging (aOR: 0.77, 95%CI:0.68-0.87). Rural regions, minor stroke/TIA as index event diagnosis, chronic renal failure, dialysis, and center type continue to have significantly reduced odds of receiving either CTA or any vascular imaging for recurrent events.

**Table 3:** Odds of vascular imaging utilization for recurrent stroke/TIA from multivariable logistic regression.

| Variable | CTA Imaging | Any Vascular Imaging |
| --- | --- | --- |
| Years Since 2015 (per year) | 1.12 (1.09-1.16)* | 1.08 (1.05-1.12)* |
| Older Patients ( ≥72 years) | 0.81 (0.72-0.92)* | 0.92 (0.81-1.05) |
| Female Sex | 0.73 (0.65-0.82)* | 0.77 (0.68-0.87)* |
| **Region** |  |  |
| Urban | Reference | Reference |
| Rural | 0.59 (0.51-0.69)* | 0.39 (0.33-0.45)* |
| **Indications** |  |  |
| Ischemic Stroke | Reference | Reference |
| Minor Stroke/TIA | 0.34 (0.30-0.39)* | 0.20 (0.18-0.23)* |
| **Comorbidities** |  |  |
| Atrial Fibrillation | 0.95 (0.83-1.10) | 0.99 (0.85-1.15) |
| Chronic Renal Failure | 0.60 (0.49-0.74)* | 0.73 (0.59-0.91)* |
| Dialysis | 0.38 (0.24-0.58)* | 0.53 (0.34-0.85)* |
| Diabetes Mellitus | 0.91 (0.81-1.03) | 0.91 (0.80-1.04) |
| Heart Failure | 0.98 (0.83-1.15) | 1.05 (0.88-1.25) |
| Hypertension | 1.09 (0.93-1.27) | 1.28 (1.09-1.51)* |
| Coronary Artery Disease | 0.91 (0.76-1.08) | 0.95 (0.79-1.15) |
| **Center Type** |  |  |
| Comprehensive Stroke Center | Reference | Reference |
| Primary Stroke Centre | 0.35 (0.30-0.40)* | 0.55 (0.47-0.66)* |
| Stroke Unit Equivalent Care | 0.08 (0.02-0.41)* | 0.10 (0.02-0.53)* |
| None of the above | 0.21 (0.18-0.24)* | 0.33 (0.28-0.39)* |

Of the 47,963 patients in our cohort, 1638 (3.4%) underwent carotid procedures. Female patients were less likely to receive carotid procedures (2.4% female vs 4.4% male, p<0.001, aOR:0.57, 95%CI:0.51-0.64). There were no significant variations in the odds of receiving carotid procedures based on year, age group, or region type. Those with minor stroke/TIA (aOR: 0.81, 95%CI:0.73-0.91) were less likely to have carotid procedures, as were those with atrial fibrillation (aOR:0.68, 95%CI:0.59-0.79). Patients with hypertension (aOR:2.19, 95%CI:1.91-2.50) or coronary artery disease (aOR:1.33, 95%CI:1.14-1.55) had increased odds of receiving carotid procedures. We did note that patients seen in primary stroke centers (aOR:0.70, 95%CI:0.62-0.79) or centers not classified as either primary, comprehensive, or stroke unit equivalent centers (aOR:0.46, 95%CI:0.39-0.53) had significantly lower odds of receiving carotid procedures compared to patients from Comprehensive Stroke Centers. Patients who had no vascular imaging (aOR:0.43, 95%CI:0.36-0.50) or ultrasound (aOR:0.25, 95%CI:0.18-0.36) were less likely to receive any carotid procedures than those who received CTA/MRA, even after accounting for all the factors above (Table 4).

**Table 4:**
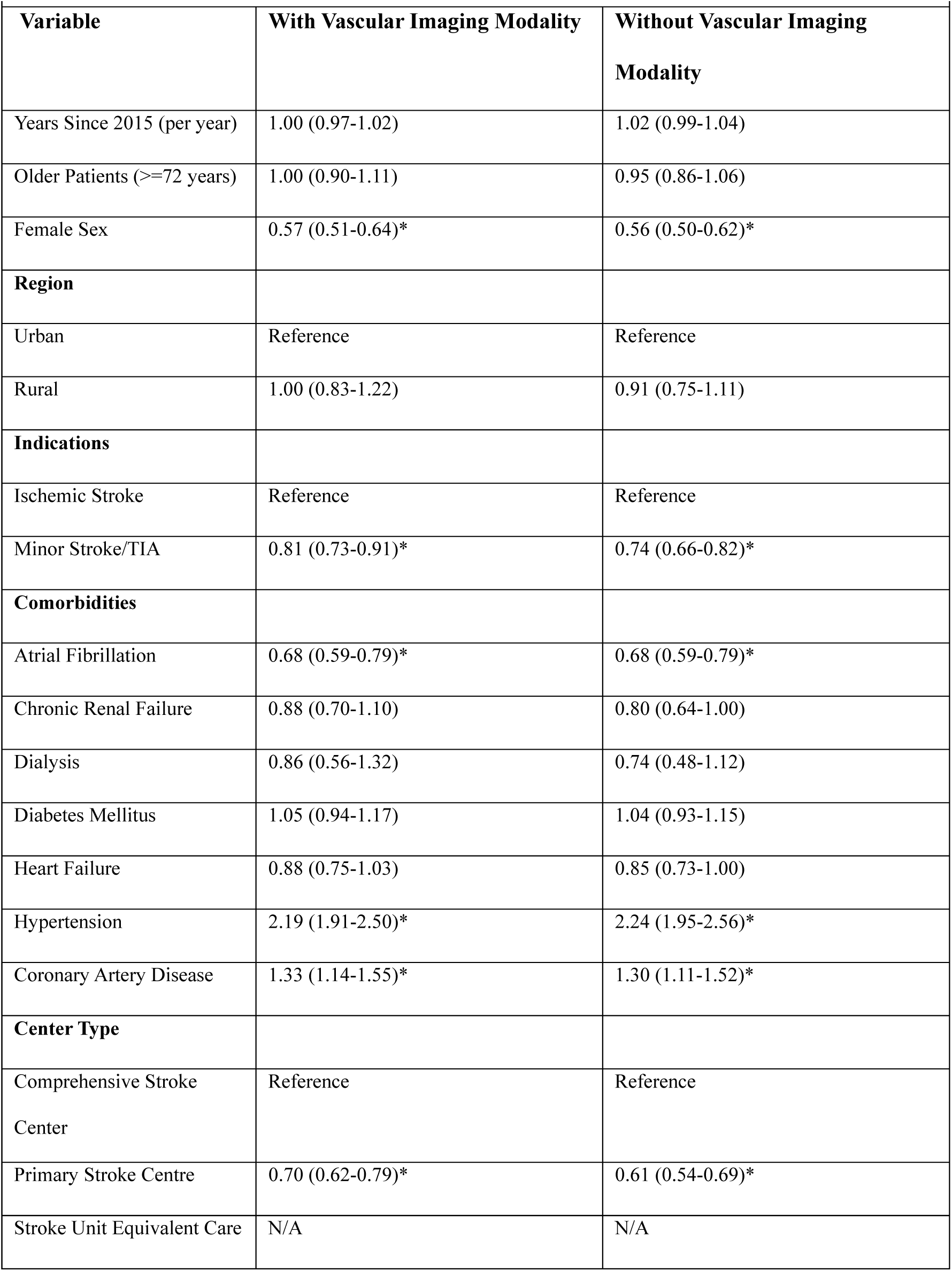
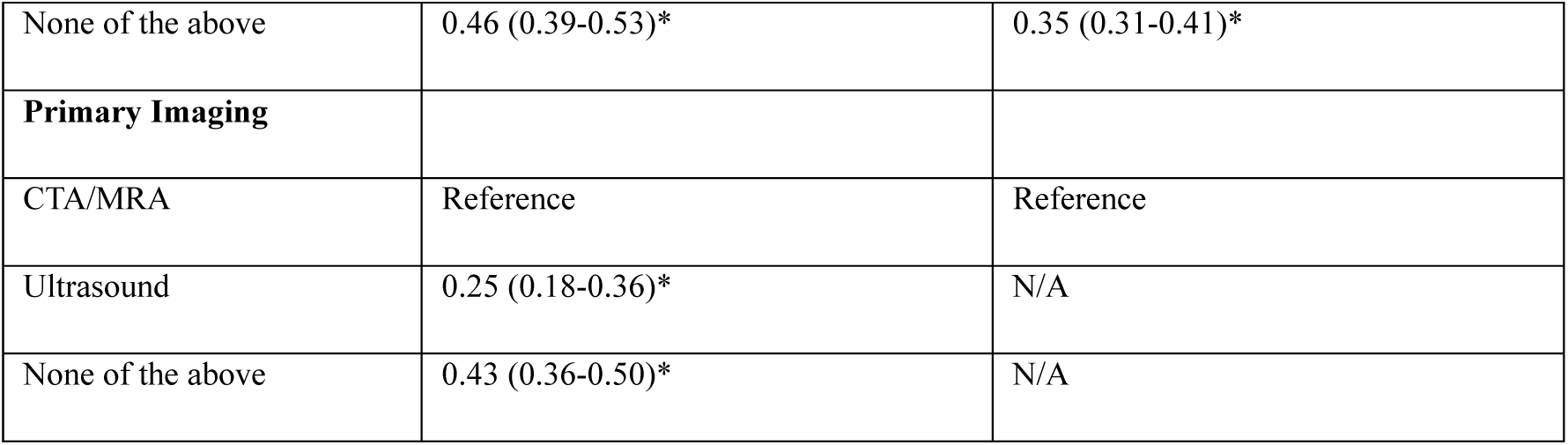
Odds of carotid revascularization procedure from multivariable logistic regression.

| Variable | With Vascular Imaging Modality | Without Vascular Imaging Modality |
| --- | --- | --- |
| Years Since 2015 (per year) | 1.00 (0.97-1.02) | 1.02 (0.99-1.04) |
| Older Patients ($\geq 72$ years) | 1.00 (0.90-1.11) | 0.95 (0.86-1.06) |
| Female Sex | 0.57 (0.51-0.64)* | 0.56 (0.50-0.62)* |
| **Region** |  |  |
| Urban | Reference | Reference |
| Rural | 1.00 (0.83-1.22) | 0.91 (0.75-1.11) |
| **Indications** |  |  |
| Ischemic Stroke | Reference | Reference |
| Minor Stroke/TIA | 0.81 (0.73-0.91)* | 0.74 (0.66-0.82)* |
| **Comorbidities** |  |  |
| Atrial Fibrillation | 0.68 (0.59-0.79)* | 0.68 (0.59-0.79)* |
| Chronic Renal Failure | 0.88 (0.70-1.10) | 0.80 (0.64-1.00) |
| Dialysis | 0.86 (0.56-1.32) | 0.74 (0.48-1.12) |
| Diabetes Mellitus | 1.05 (0.94-1.17) | 1.04 (0.93-1.15) |
| Heart Failure | 0.88 (0.75-1.03) | 0.85 (0.73-1.00) |
| Hypertension | 2.19 (1.91-2.50)* | 2.24 (1.95-2.56)* |
| Coronary Artery Disease | 1.33 (1.14-1.55)* | 1.30 (1.11-1.52)* |
| **Center Type** |  |  |
| Comprehensive Stroke Center | Reference | Reference |
| Primary Stroke Centre | 0.70 (0.62-0.79)* | 0.61 (0.54-0.69)* |
| Stroke Unit Equivalent Care | N/A | N/A |
| None of the above | 0.46 (0.39-0.53)* | 0.35 (0.31-0.41)* |
| **Primary Imaging** |  |  |
| CTA/MRA | Reference | Reference |
| Ultrasound | 0.25 (0.18-0.36)* | N/A |
| None of the above | 0.43 (0.36-0.50)* | N/A |

## Discussion

Using population-level data from an entire Canadian province, we found that 60.7% of the patients with ischemic stroke or minor stroke/TIA received CTA, with increased utilization over the years, ranging from 49.1% in 2016 to 77.9% in 2023. We also note a prominent reduction in the percentage of individuals receiving no vascular imaging, from 40.5% of patients in 2016 to 14.0% in 2023. This highlights the increased uptake of appropriate vascular imaging, possibly due to the increasing use carotid procedures and better adherence to guidelines.

One of the most concerning findings from this study was the significantly lower odds of female patients receiving CTA or any vascular imaging compared to male patients. Previous studies have reported female patients receiving brain imaging and angiography less frequently than male counterparts^15^. Although certain situations, such as pregnancy, may make clinicians concerned about the fetal risk of radiation exposure, current guidelines highlight that CTA may still be used given the negligible risks of the radiation and there is no known harm caused to the fetus by CT contrast dye^16^. Our analysis also revealed female patients had lower odds of receiving carotid procedures; this may potentially relate to the concern that female patients have more surgical risks and reduced benefits than male patients. Although a systematic review from the early 2000s did conclude female patients had higher rates of operative stroke and death when undergoing carotid endarterectomy, multiple other studies have shown no difference in complication rates or benefits^17–20^.

We also found that compared to patients presenting to hospitals in urban regions, patients in rural sites were less likely to receive CTAs or any vascular imaging. This may be attributed to rural sites’ limited diagnostic imaging capabilities and access to specialist care^21^; however, integrated stroke systems of care should strive to ensure that patients initially seen at such sites are still connected to appropriate imaging through other facilities in the system. We also found patients presenting to centers other than CSCs (the latter present only in two large urban centers in our province) were less likely to receive vascular imaging, including CTA. Further, our findings revealed that the failure to receive any vascular imaging was associated with lower odds of carotid procedure and that patients presenting to non-CSC facilities had lower odds of receiving carotid procedures even after adjusting for imaging received. Previous literature has reported regional variation in carotid revascularization; however, we did not specifically find any significant disparities between urban vs rural centers after taking into account center type^10,22^.

Our analysis also found that older patients were less likely to receive CTA or any vascular imaging. Vascular imaging is pivotal in this demographic, as previous studies have illustrated that carotid procedures in elderly patients are safe and provide significant benefits^23,24^. We also found significantly lower odds of receiving CTA or any vascular imaging in patients with index diagnosis of minor stroke/TIA. Although this may be due to the milder and transient nature of these presentations, international stroke guidelines emphasize that all patients with acute disabling or non-disabling stroke, as well as TIAs, should undergo neurovascular imaging to identify carotid stenosis; in the Canadian guidelines, CTA is recommended as the first-line imaging, although MRA and ultrasound are mentioned as appropriate alternatives^25–27^. Finally, patients with specific comorbidities, namely chronic renal failure, were less likely to receive CTAs. Although clinicians may be concerned about using intravenous contrast in patients with known renal disease due to the risk of contrast-induced nephropathy, either ultrasound or MRA may be used in lieu; however, our results reveal that patients with these comorbidities are less likely to receive any modality of non-invasive vascular imaging. Further, it is important to emphasize that imaging-associated renal complications are rare, and the benefits of prompt imaging often outweigh the risks^28,29^.

There are several limitations to our study. Patient ethnicity and/or race were not collected in our administrative data. We did not have information about stroke etiology or the presence of significant carotid disease on imaging, both of which would influence decisions to pursue revascularization. However, it would, of course, be impossible to know the carotid disease status for the patients who did not receive vascular imaging in the first place, and it was essential for us to include these patients as well in the analysis to understand disparities at a population level. We also did not have data on whether patients were offered carotid procedures or vascular imaging but refused them. We did not specifically quantify stroke severity beyond diagnostic classification into major stroke, minor stroke, or TIA. However, it is essential to note that neurovascular imaging, especially CTA, is pivotal even for TIAs and minor strokes to establish stroke etiology and guide management for secondary prevention^30^. As we examined patients presenting with stroke or TIAs in Alberta, our findings may not be generalizable to other regions with varying health resources and systems; however, as a population study, the results have provided us invaluable insights into disparities in stroke imaging and subsequent carotid procedures.

In conclusion, we found disparities in the utilization of vascular imaging based on whether the hospital was in an urban or rural region, further modulated by patient characteristics, such as sex, age, and center type, despite increasing utilization over time. We discovered that female patients had significantly lower odds of vascular imaging and carotid procedures. Further studies must investigate the morbidity and mortality endpoints due to these disparities and analyze contributory factors that may be addressed to improve patient health outcomes.

## Data Availability

Requests for the data will be considered by the corresponding author when accompanied by an appropriate scientific proposal

## Non-standard Abbreviations and Acronyms

TIA: Transient ischemic attack
MRA: Magnetic resonance angiography
CTA: Computed tomography angiography
CSC: Comprehensive stroke center
PSC: Primary stroke center
SUEC: Stroke-unit equivalent center

